# Effect of combination of interferon alpha-2b and interferon-gamma or interferon alpha-2b alone for elimination of SARS-CoV-2 viral RNA. Preliminary results of a randomized controlled clinical trial

**DOI:** 10.1101/2020.07.29.20164251

**Authors:** Esquivel-Moynelo I Idelsis, Pérez-Escribano J, Duncan-Roberts Y, Vazquez-Blonquist D Dania, Bequet-Romero M, Baez-Rodríguez L, Castro-Ríos J, Cobas Cervantes L, Pagé-Calvet E, Travieso-Pérez S, Martinez-Suarez C, Campa-Legra I Ivan, Fernandez-Masso Julio Raul, Camacho-Rodriguez H, Díaz-Gálvez M, Sin-Mayor A, García-Sánchez M, Martínez-Martín SM, Alonso-Valdés M, Hernandez-Bernal F, Nodarse-Cuni H, Bello-Garcia D, Beato-Canfuk A, Vizcaino M Tania, Guillen-Nieto GE, Lucila Muzio-Gonzalez VL Verena, Bello-Rivero I

## Abstract

**Objectives:** An IFN-α2b and IFN-γ combination has demonstrated favorable pharmacodynamics for genes underlying antiviral activity which might be involved in the defense of a host from a SARS-CoV-2 infection. Considering this synergy, we conducted a randomized controlled clinical trial for efficacy and safety evaluation of subcutaneous IFN - α2b and IFN-γ administration in patients positive for SARS-CoV-2.

**Methods:** We enrolled 19-82 years-old inpatients at the Military Central Hospital Luis Diaz Soto, Havana, Cuba. They were hospitalized after confirmed diagnosis for SARS-CoV-2 RNA by real-time reverse transcription polymerase chain reaction. Patients were randomly assigned in a 1:1 ratio to receive either, subcutaneous treatment with a co-lyophilized combination of 3.0 MIU IFN-α2b and 0.5 MIU IFN-γ (HeberFERON, CIGB, Havana, Cuba), twice a week for two weeks, or thrice a week intramuscular injection of 3.0 MIU IFN-α2b (Heberon® Alpha R, CIGB, Havana, Cuba). Additionally, all patients received lopinavir-ritonavir (200/50 mg every 12 h) and chloroquine (250 mg every 12 h, i.e.standard of care). The primary endpoints were, from the start of treatment, the time to elimination of viral RNA and the time to progression to severe COVID-19. The protocol was approved by the Ethics Committee on Clinical Investigation from the Hospital and the Center for the State Control of Medicines, Equipment and Medical Devices in Cuba. Informed consent was obtained from each participant (INSTITUTION PROTOCOL IG/IAG/CV/2001).

**Results:** A total of 79 patients with laboratory-confirmed SARS-CoV-2 infection, including symptomatic or asymptomatic conditions, fulfilled the inclusion criteria and underwent randomization. Thirty-three subjects were assigned to the HeberFERON group, and 33 to the Heberon Alpha R group. Sixty-three patients were analyzed for viral elimination, of these 78.6% in the HeberFERON group eliminated the virus after 4 days of treatment versus 40.6% of patients in the Heberon Alpha R groups (p=0.004). Time to reach the elimination of SARS-CoV-2, as measured by RT-PCR was 3.0 and 5.0 days for the HeberFERON and Heberon Alpha R groups, respectively. A significant improvement in the reduction of time for virus elimination was attributable to HeberFERON (p=0.0027, Log-rank test) with a Hazard Ratio of 3.2 and 95% CI of 1.529 to 6.948, as compared to the Heberon Alpha R treated group.

Worsening of respiratory symptoms was detected in two (6.6%) and one (3.3%) patients in HeberFERON and IFN-α2b groups, respectively. However, none of the subjects transited to severe COVID-19 during the study or during the following clinical evaluation (21 more days).

RT-PCR on day 14 after the start of the treatment was negative to SARS-CoV-2 in 100% and 91% of patients of the combination of IFNs and IFN-α2b, respectively. Elimination in HeberFERON treated patients was related to a significant increase in lymphocytes counts and also a significant reduction in CRP as early as 7 days after commencing the therapeutic schedule.

All the patients in both cohorts recovered and had their laboratory parameters return to normal values by day 14 after treatment initiation. Adverse events were identified in 31.5% of patients, 28.5% in the control group, and 34.4% in the HeberFERON group, with the most frequent adverse event being headaches (17.4%).

**Conclusions:** In a cohort of 63 hospitalized patients between 19 to 82 years-old with positive SARS-CoV-2, HeberFERON significantly eliminated the virus on day 4 of treatment when compared to treatment with IFN-α2b alone. However, Heberon Alpha R alone also showed efficacy for the treatment of the viral infection. Both treatments were safe and positively impacted on the resolution of the symptoms. None of the patients developed severe COVID-19.

## Introduction

Coronavirus disease (COVID-19) is an infectious disease caused by SARS-CoV-2 that has spread to more than 230 countries of the World and generated more than 644 832 deaths, with great social and economic consequences. Since effective vaccines are not available, it is urgent to find and develop strategies to ameliorate this virus effect^1^. The first cases of the disease in Cuba were confirmed on March 11, 2020: three tourists from the Italian region of Lombardy, who were immediately hospitalized^2^.

In Cuba, 60.1% of people diagnosed with COVID-19 are less than 20-years-old. In the country 53.9% of patients are asymptomatic, and even in the most vulnerable population of patients, comprising the 80-year-old group, 51.7% of those infected present with no symptoms at virus confirmation^3^.

Due to its antiviral nature, interferons (IFNs) have been used for the treatment of viral infections. Their therapeutic use is justified by the antiviral and immunomodulatory properties of these proteins^4^. In fact, severity of COVID-19 disease correlates with the failure to initiate an IFN response to SARS-CoV-2 infection^5^.

Taking these into consideration and the fact that therapeutics that target the coronavirus alone, might not be able to reverse highly pathogenic infections, the Cuban Protocol for Management of COVID-19^2^ includes Heberon Alpha R and other antiviral treatments for the symptomatic phase. Cuban patients already showing symptoms or their near contacts are isolated in centers established for that purpose and start receiving treatment. As mentioned, this schedule incorporates Heberon Alpha R with lopinavir-ritonavir (Kaletra) and chloroquine (CQ). After confirmation of a positive SARS-CoV-2 test, they are hospitalized and continue to, or start to receive Heberon Alpha R, Kaletra, and CQ as established by Cuban Health Ministry guide-lines^2^.

This has resulted in a favorable evolution of the patients in a cohort of 761 subjects confirmed for SARS-CoV-2 infection and receiving Heberon Alpha R, where 95.4% fully recovered from COVID-19, with only a 0.92% case fatality rate^6^.

Earlier studies of a combination of type I IFN and IFN-γ shown a synergistic inhibition of the SARSCoV virus replication in vitro^7,8,9,10^. IFN-γ is a key moderator in linking the innate immunity to adaptive immune responses^11^, hence it is possible that a combinational therapy of IFNs and other antiviral drugs could significantly inhibit virus replication and modulate clinical variables related to the immune response, with a positive outcome in terms of viral infection resolution^12,13,14^.

In accordance with the previous, we conducted a phase 2 randomized trial to establish whether a combination of IFN-α2b and IFN-γ with the standard of care, can improve the viral load profile and clinical parameters in adults with COVID-19.

## Methods

### Study design

Hospitalized adult patients with RT-PCR confirmed SARS-CoV-2 were enrolled in this open-labeled, single center, prospective, randomized and controlled clinical trial at Military Central Hospital Luis Diaz Soto Hospital, Havana, Cuba.

Patients were randomly assigned to receive the combination of IFN-α2b and IFN-γ (HeberFERON, CIGB, Havana, Cuba) or IFN-α2b (Heberon Alpha R, CIGB, Havana, Cuba) based on a power of 80%, and a level of confidence set at 95%, while also considering a dropout rate of 5%. Patients were randomized individually to one of two treatment arms by means of random computer-generated lists, with an allocation ratio of 1:1, with block sizes of six patients.

Heberon Alpha R (IFN-α2b) is a drug produced in Cuba by the Center for Genetic Engineering and Biotechnology (CIGB), which has remained a product with proven antiviral efficacy and an adequate safety profile for 34 years^15^. HeberFERON (IFN-α2b and IFN-γ, co-lyophilized in the same vial) is produced at CIGB, and registered in Cuba for the treatment of basal cell carcinoma.^16^

The study execution followed the ethical principles of the Declaration of Helsinki and the International Council for Harmonization–Good Clinical Practice guidelines. No compensation was provided for enrollment in the trial. Patient personal data was protected. The authors were responsible for designing the trial and for collecting and analyzing the data. The authors assured the completeness and accuracy of the data collection and the adherence to the protocol. The details of the trial are provided in the protocol that has been posted in TRIALS^17^ and is in processing by the editors of the journal.

The primary endpoints were the time to viral RNA elimination from the start of treatment and the time to progression to severe COVID-19.

### Eligibility criteria

The COVID-19 diagnosis was obtained by a positive real-time reverse transcription-polymerase chain reaction (RT-PCR) amplification of the viral E gene and then confirmation by amplification of the RdRP gene from throat swab samples. Adult (≥19 years-old) patients with RT-PCR confirmed SARS-CoV-2, ECOG functional status ≥ 2 (Karnofsky ≥ 70%), and who volunteered by signing the informed consent were included. Patients with each of the following characteristics were excluded: decompensated chronic diseases at the time of inclusion (severe arterial hypertension, ischemic heart disease, diabetes mellitus, etc.), with a history of autoimmune diseases, presence of hyper inflammation syndrome, serious coagulation disorders, known hypersensitivity to any of the components of the formulation under evaluation, pregnancy or lactation, and obvious mental incapacity to issue consent and act accordingly with the study.

The clinical trial protocol was approved by the Ethics Committee on Clinical Investigation of Military Central Hospital Luis Diaz Soto, and the Center for the State Control of Medicines, Equipment and Medical Devices (CECMED) in Cuba. Patients were asked for written consent to participate after having been duly informed about the characteristics of the trial, objectives, benefits and possible risks. Likewise, they were informed of their rights to participate or not and to withdraw their consent at any time, without exposing themselves to limitations for their medical care or other retaliation. The study was registered on April 2020 at: registroclinico.sld.cu/en/trials/RPCEC00000307.

After a preliminary exploratory analysis of the outcomes of the first 79 patients, the monitoring board considered a preliminary report and early publication of the RT-PCR results from the 63 patients with available throat swabs, due to the significant effect of HeberFERON on the reduction of the time to viral clearance. The trial finally included 134 patients from whom samples are now in the process of data collection for definitive processes and analysis.

### Treatment protocols

Patients received 3.0 million international units (MIU) IFN-α2b and 0.5 MIU IFN-γ (HeberFERON), twice a week for two weeks, subcutaneously and lopinavir-ritonavir 200/50 mg every 12 h and CQ 250 mg every 12 h (treatment group); or standard of care (3.0 MIU IFN-α2b (Heberon Alpha R), thrice a week, intramuscularly and lopinavir-ritonavir 200/50 mg every 12 h and CQ 250 mg every 12 h (control group).

### Data collection

Demographic, clinical, laboratory, treatments and outcome characteristics of patients were extracted from medical records and registered in CRF and then were entered in duplicate (independently by two operators) for the subsequent process of automatic comparison and correction of the databases, necessary for statistical analysis with accurate information from the trial. However the blinding was not feasible, it was maintained for laboratory SARS-CoV-2 RNA detection by RT-PCR, one of the endpoints of the study.

### Laboratory procedures

The hospital received patients from several zones in Havana city diagnosed in reference centers for SARS-CoV-2 infection following the Cuban Ministry of Health guidelines for diagnostic testing. Patients were defined to have SARS-CoV-2 if they had two consecutive positive results, including the confirmatory test by RT-PCR targeting amplifications of E and /or RdRP genes. A cycle threshold up to value 40 was defined as positive.

Specimens were obtained from throat swabs of patients at the hospital following standard procedures and transported to a BSL2 certified laboratory at the CIGB for serial evaluation of SARS-CoV-2 viral nucleic acid detection by RT-PCR after extraction utilizing the QIAamp® Viral RNA Mini kit (Qiagen, USA). A multiplexed detection by RT-PCR was carried out targeting E and/or RdPR genes plus an EAV internal extraction control (TIB MOLBIOL Syntheselabor GmbH, Berlin, Germany) as described using the Multiplex RNA Virus Master (Roche, USA).

Routine blood examinations were done at the Clinical Laboratory of Military Central Hospital Luis Diaz Soto and included a whole blood count, coagulation profile, serum biochemical tests (including renal and liver function, electrolytes, and coagulation), C-reactive protein (induced by various inflammatory mediators such as IL-6^18^), and ferritin. Furthermore, all patients received a chest X-ray. The frequency of examinations were defined in the trial protocol and consisted of weekly determinations at base line on days 2 and 4 of each week.

### Outcomes assessment

The primary outcomes included virological and clinical evaluations. Time to SARS-CoV-2 RNA elimination (absence of the virus according to the RT-PCR) in positive patients after starting antiviral therapy was the viral endpoint. It was expressed as the percentage of patients negative for SARS-CoV-2 by RT-PCR in throat exudate tissue extracts calculated at 48, 72, 96 and 120 hours. Days to viral elimination were analyzed by Kaplan-Meier plots and compared with a Log-rank test. Hazard Ratio with 95% CI was also calculated.

The clinical evaluation considered the time to progression to severe COVID-19 and it was calculated by the percentage of patients who became severely ill after the end of the antiviral treatment under investigation.

### Statistical analysis

Quantitative variables were described with the arithmetic mean and its standard deviation and the median with its range. We used the absolute and relative frequency (%) for qualitative variables. The hypothesis test used was Fisher’s exact test. The viral elimination analysis was performed using the Kaplan-Meier plot representation and the comparison of factors was done with the Mantel-Cox Log Rank tests. The evolution of laboratory parameters while under treatment was analyzed using a paired mixed model (which cannot handle missing values appearing due to patient release from hospital). Correlations between virus elimination and laboratory parameters were studied using a two-tailed non-parametric Spearman correlation with 95% confidence interval. P<0.05 was considered statistically significant. Statistical analysis was performed using the Windows software package SPSS (version 25) and GraphPad Prism v8.0.

## Results

### Patients and baseline features

We screened 144 patients positive by RT-PCR for SARS-CoV-2. Fifty-seven patients did not fulfill the inclusion criteria, i.e. one with icterus, one with chronic decompensate renal insufficiency, two non-confirmed positive PCR for SARS-CoV-2, and fifty-three patients with 2 positive RT-PCR after more than 21 days of persistent virus shedding. Patients with viral persistence were later treated with the HeberFERON out of the clinical trial (manuscript in preparation). Eight patients that did not consent were also excluded.

Seventy-nine subjects met the inclusion criteria and were randomly assigned (1:1) to either the HeberFERON group (41 patients) or the control group (38 patients). Twelve patients did not start the treatment, 7 patients refused to start the treatment, although they signed the consent, and 5 were excluded due to loss of inclusion criteria.

Seven patients withdrew due to several causes: 3 due to worsening of respiratory symptoms (two of them in the HeberFERON group, with asthma as the underlying disease) that changed to the other exclusions in the study drug treatment; 3 patients from the control group with positive RT-PCR on day 14 were switched to receive HeberFERON outside of the clinical trial by medical decision; and 1 with the appearance of an exclusion criterion (pregnancy) in the control group (see figure 1, flow chart of the study).

**Figure 1.**
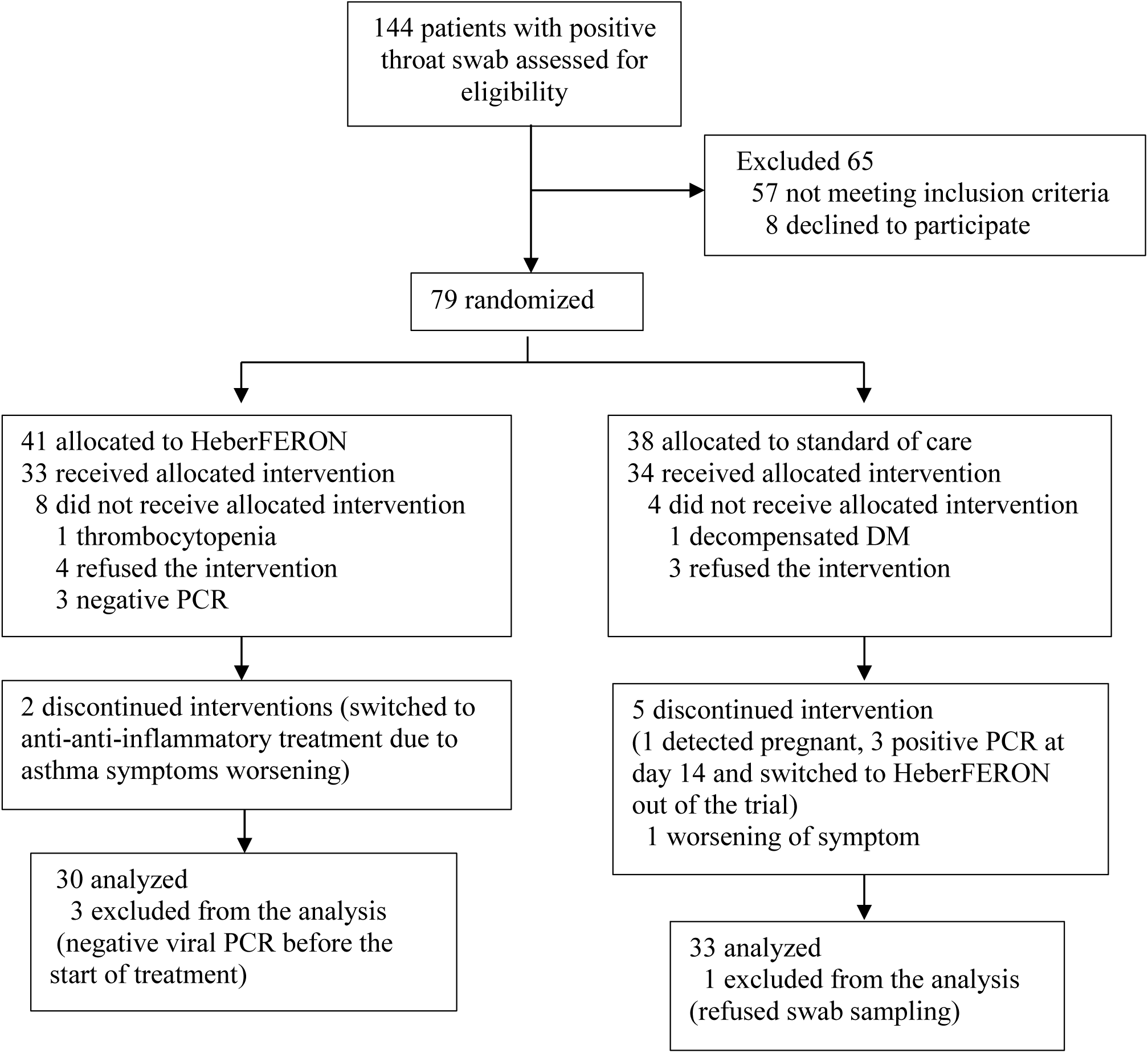
Randomization and treatment assignment.

Four patients were not analyzed, three in the HeberFERON group, due to bad inclusions (negative for viral RNA before the beginning of treatment as identified by the board of monitors), and one in the control group who refused the swabs sampling. Thirty and thirty-three patients were analyzed by intention to treat (ITT) in the HeberFERON and control group, respectively.

Finally, swabs samples to test for viral elimination were obtained from sixty-three patients. In this cohort 29 were symptomatic (46.0%), with a median from the beginning of symptoms to the start of treatments with IFNs of 7.0 days (IQR: 2-13) in the control group and 7.5 days (IQR: 2-19) in the HeberFERON group.

Thirty-three patients were treated with standard of care which includes Heberon Alpha R (control group) and 30 with HeberFERON plus standard of care, excluding Heberon Alpha R. In the control group younger people prevailed with a median of 31.0 years-old (IQR: 19-57), while those 42.0 years-old (IQR: 19-82) were in the HeberFERON group (p=0.023). The sex distribution showed a prevalence of males in the control cohort (20/33, 60.6%) as compared with a similar distribution in the combination group for males (14/30, 46.7%) and females (see table 1); however this difference was not statistical significant.

**Table 1.** Demographic and Clinical Characteristics of the Participants at Baseline.

| **Characteristic** | **HeberFERON**<br>**(n=30)** | **Control group**<br>**(n=33)** | **Total**<br>**(n=63)** |
| --- | --- | --- | --- |
| **Median age (IQR) —yr ¶** | 42.0 (19-82) | 31 (19-57) | 38 (19-82) |
| **Sex** |  |  |  |
| Male — no. (%) | 14 (46.7) | 20 (60.6) | 34 (54.0) |
| Female — no. (%) | 16 (53.3) | 13 (39.4) | 29 (46.0) |
| **Current smoker— no. (%)** | 5 (15.1) | 3 (9.0) | 8 (12.7) |
| **Symptomatic— no. (%)** | 12 (40.0) | 17 (51.5) | 29 (46.0) |
| Median age (IQR) —yr | 50 (19-80) | 23 (19-57) | 43 (19-80) |
| Median time (IQR) from symptom onset to randomization — days | 7.5 (2-19) | 7 (2-13) | 7 (2-19) |
| Male — no. /total no. (%) § | 4/12 (33.3) | 11/17 (70.6) | 15/29 (51.7) |
| Female — no. /total no. (%) | 8/12 (66.6) | 5/17 (29.4) | 13/29 (44.8) |
| **Time from symptoms onset to start of treatment, days— no./total no. (%)** |  |  |  |
| >7 days | 6/12 (50.0) | 6/16 (37.5) | 12/28 (42.9) |
| ≤7 days | 6/12 (50.0) | 10/16 (62.5) | 16/28 (57.1) |
| **Asymptomatic— no. (%)** | 18 (60.0) | 16 (48.5) | 34 (54.0) |
| Median age (IQR) —yr | 36.5 (23-82) | 34 (19-50) | 34 (19-82) |
| Male — no. /total no. (%) § | 10/18 (55.5) | 9/16 (56.2) | 19/34 (55.8) |
| Female — no. /total no. (%) | 8/18 (44.4) | 8/16 (50%) | 16/34 (47.0) |
| **Coexisting conditions— no. (%)** |  |  |  |
| Any comorbidities | 19 (57.5) | 13 (39.3) | 32 (50.8) |
| Cardiac diseases | 2 (6.0) | 2 (6.0) | 4 (6.3) |
| Diabetes | 3 (9.0) | 0 (0.0) | 3 (4.7) |
| Hypertension | 7 (21.2) | 7 (21.2) | 14 (22.2) |
| Asthma | 2 (6.0) | 2 (6.0) | 4 (6.3) |
| **Vital signs** |  |  |  |
| Median vital signs (IQR) |  |  |  |
| Temperature (°C) | 36.0 (36-37.1) | 36.2 (36-37.9) |  |
| Respiratory rate (breaths/minute) | 19 (15-22) | 19 (16-24) |  |
| Systolic blood pressure (mmHg) | 120 (100-170) | 120 (90-140) |  |
| **Baseline Treatment— no. (%)** |  |  |  |
| IFNα-2b | 30 (100%) | 33 (100%) | 63 (100%) |
| Lopinavir-ritonavir at baseline | 30 (100%) | 33 (100%) | 63 (100%) |
| Chloroquine at baseline | 30 (100%) | 33 (100%) | 63 (100%) |
| Antibiotic treatment at baseline | 1 (3.0) | 4 (12.1) | 5 (7.9) |
| **Laboratory baseline data** | **n=27** | **n=30** | **n=57** |
| Median laboratory values (IQR) |  |  |  |
| White blood cell count (**WBC**), ×10<sup>9</sup> per L | 6.4 (3.8-11.7) | 6 (2.4-12) | 6 (2.4-11.7) |
| Lymphocyte count, × 10<sup>9</sup> per L | 2.45 (0.5-3.93) | 2.3 (0.91-3.7) | 2.45 (0.5-3.93) |
| Lymphocyte Percentage (%) | 38 (11-57) | 40 (8-72) | 39 (8-72) |
| Neutrophils count, 10<sup>9</sup> per L | 3.2 (1.42-8.66) | 2.8 (0.62-9.9) | 2.92(0.62-9.86) |
| Neutrophils Percentage (%) | 50 (29-81) | 48 (18-85) | 49 (18-85) |
| Platelet count, × 10<sup>9</sup> per L | 224 (117-351) | 235 (126-376) | 226 (117-376) |
| Neutrophils to Lymphocyte Ratio (**NLR**) | 1.34 (0.51-6.82) | 1.2 (0.25-11) | 1.25 (0.25-10.6) |
| Platelet to Lymphocyte Ratio (**PLR**) | 107.3 (34-327.7) | 103 (62-320) | 107.3(34-327.7) |
| Systemic Immune inflammation Index<br>(SII) | 306 (115-2075) | 275 (73-2879) | 289 (72.5-2869) |
| Serum creatinine, $\mu\text{mol/L}$ | 99.6 (31.6-155) | 103 (77-143) | 102.4 (31.6-155) |
| Aspartate aminotransferase, U/L | 28.1 (18.9-83) | 26 (14-97) | 25.7 (13.6-96.70) |
| Alanine aminotransferase, U/L | 34.05 (17.3-133.2) | 35 (14-78) | 34.7 (13.8-133.2) |
| Creatine kinase, U/L | 113 (31.09-2377) | 110 (81-143) | 109 (31.09-237) |
| C-Reactive Protein mg/dL | 4.135 (0.38-129) | 2 (0.29-37) | 3.58 (0.29-129) |
| Ferritin ng/mL | 206 (40.7-873) | 190 (12-600) | 197 (11.6-10.2) |
| TP seg | 13.95 (11-16.6) | 13 (12-16) | 13.5 (11-16.6) |
| **Ionogram** |  |  |  |
| Potassium mmol/L | 103 (95.5-113) | 100 (96-109) | 103 (92-112) |
| Sodium mmol/L | 143 (136-151) | 142 (136-147) | 142 (134-151) |
| Chloride mmol/L | 4.1 (2.66-5.4) | 4.1 (2.9-5.1) | 4.04 (2.66-5.4) |

p=0.023 by U de Mann-Whitney; \$ p= 0.024 for the comparison between sex S and A by the Chi-square.

In the control group more symptomatic patients were present (51.5%) than in the HeberFERON group (40.0%); however this difference was not statistically significant.

The median age of symptomatic subjects was higher in the HeberFERON group [50 (IQR: 19-80)] than in the control (24 [IQR: 19-57)]. In the HeberFERON group, 66.6% of symptomatic were females and in the control group symptomatic males were more frequent 70.6% (p=0.024). Symptomatic patients with more than 7 days from symptoms onset were more common in the HeberFERON arm (50.0%). However, these numerically differences were not statistically significant (see table 1).

The more common symptoms (table 1) were fever and unproductive cough (16.4%), followed by headache (9.6%), decay (8.4%), odynophagia and nasal secretions (5.4%), diarrhea, dyspnea, chills and general malaise (4.1%), and sore throat and myalgia (2.7%).

Fifty percent of patients had any comorbidity; the most frequent were hypertension (22%), asthma (6.3%), diabetes and glaucoma (4.7%).

The vital signs at the time of hospital admission were not statistically different between groups.

Some imbalances existed at enrollment between the groups, including a higher median age in the HeberFERON than in the control group, as well as more patients with higher than 7 days from onset of the symptoms in the HeberFERON group.

No other major differences in symptoms, vital signs, laboratory results, disease severity, or treatments were observed between groups at baseline.

### Outcomes

We analyzed 63 patients with available throat swab samples. In the HeberFERON group 78.6% of patients were negative for the virus after 4 days of treatment versus 40.6% of patients in the control group (p=0.004). HeberFERON eliminated SARS-CoV-2 in the 95.8% of patients at day 5, (p=0.0479), see tables 2 and 3.

**Table 2.** Outcomes in patients positive for SARS-CoV-2 and treated with interferons.

| Outcomes |  | HeberFERON<br>(n=30) | Control group<br>(n=33) | p |  |
| --- | --- | --- | --- | --- | --- |
| Time to reach 50% the negativization of the SARS-CoV-2 (Log-rank test) | All patients | 3 days | 5 days | 0.002 |  |
|  | Symptomatic | 2.5 days | Not reached | 0.003 |  |
|  | Asymptomatic | 3 days | 5 days | 0.123 |  |
| Percent of patients negative to viral RNA after the start of treatment (Fisher test) | 48 h | 44.8 | 15.4 | 0.022 |  |
|  | 72 h | 63.3 | 34.5 | 0.037 |  |
|  | 96 h | 78.6 | 40.6 | 0.004 |  |
|  | 120 h | 95.8 | 73.9 | 0.047 |  |
| Time to severe COVID-19 |  | Non-reached | Non-reached |  |  |
| Percent of patients with clinical symptom worsening |  | 6.66% | 3.33% |  |  |
| Percent of hospital discharges at day 14 |  | 100% | 91% |  |  |
| Frequency of adverse events |  | 34.4% | 28.5% |  |  |
| Correlation studies of laboratory parameters and time to negative viral RNA PCR results |  |  |  |  |  |
| Parameters |  | 7 days on HeberFERON |  | 7 days on control group |  |
|  |  | r | p | r | p |
| Lymphocyte Concentration | All patients | -0.5220 | **0.0336** | -0.0039 | 0.9849 |
|  | Symptomatic | -0.7373 | **0.0197** | 0.2154 | 0.4768 |
| C-Reactive Protein (CRP) | All patients | 0.5016 | **0.0439** | -0.2393 | 0.5822 |
|  | Symptomatic | 0.5339 | 0.1160 | -0.5372 | 0.0610 |
| Creatinine Phosphokinase Kinase (CPK) | All patients | -0.020 | 0.9392 | 0.4097 | **0.0304** |
|  | Symptomatic | -0.1653 | 0.6470 | -0.3840 | 0.1749 |
| Platelet to Lymphocyte Ratio (PLR)_ | All patients | 0.4978 | **0.0439** | 0.02942 | 0.8866 |
|  | Symptomatic | 0.7373 | **0.0197** | -0.2882 | 0.3371 |

**Table 3.** Detection of SARS-CoV-2 in swabs by RT-PCR in the HeberFERON or control groups. Throat swabs were taken from 63 COVID-19 positive patients at 48h, 72h, 96h and 120h after their treatment with HeberFERON (30 patients) or standard of care (33 patients). Viral nucleic acid detection was carried out by RT-PCR resulting in positive or negative samples. For each analysis time, we represent the number of positive and negative patients, the percentage of virus elimination and the p value in a Fisher test analysis in an overall analysis (ALL) and dividing patients as Symptomatic (S) and Asymptomatic (A). *: p< 0.05; **: p> 0.01; ns: p> 0.05.

|  |  |  | HeberFERON | Control group |
| --- | --- | --- | --- | --- |
| 48h | ALL | Positives | 16 | 22 |
|  |  | Negatives | 13 | 4 |
|  |  | No Data | 1 | 7 |
|  |  | % virus elimination | 44.8 | 15.4 |
|  |  | p (Fisher test ) | 0.0224 (*) |  |
|  | S | Positives | 5 | 11 |
|  |  | Negatives | 6 | 2 |
|  |  | No Data | 1 | 3 |
|  |  | % virus elimination | 54.5 | 15.4 |
|  |  | p (Fisher test ) | 0.0825 (ns) |  |
|  | A | Positives | 11 | 10 |
|  |  | Negatives | 7 | 2 |
|  |  | No Data |  | 4 |
|  |  | % virus elimination | 38.9 | 16.7 |
|  |  | p (Fisher test ) | 0.2487 (ns) |  |
| 72h | ALL | Positives | 11 | 19 |
|  |  | Negatives | 19 | 10 |
|  |  | No Data |  | 4 |
|  |  | % virus elimination | 63.3 | 34.5 |
|  |  | p (Fisher test ) | 0.0379 (*) |  |
|  | S | Positives | 3 | 9 |
|  |  | Negatives | 9 | 5 |
|  |  | No Data |  | 2 |
|  |  | % virus elimination | 75.0 | 35.7 |
|  |  | p (Fisher test ) | 0.0618 (ns) |  |
|  | A | Positives | 8 | 9 |
|  |  | Negatives | 10 | 5 |
|  |  | No Data |  | 2 |
|  |  | % virus elimination | 55.6 | 35.7 |
|  |  | p (Fisher test ) | 0.3075 (ns) |  |
| 96h | ALL | Positives | 6 | 19 |
|  |  | Negatives | 23 | 13 |
|  |  | No Data | 1 | 1 |
|  |  | % virus elimination | 78.6 | 40.6 |
|  |  | p (Fisher test ) | 0.004 (**) |  |
|  | S | Positives | 1 | 10 |
|  |  | Negatives | 11 | 6 |
|  |  | No Data |  |  |
|  |  | % virus elimination | 91.7 | 37.5 |
|  |  | p (Fisher test ) | 0.0060 (**) |  |
|  | A | Positives | 5 | 8 |
|  |  | Negatives | 12 | 7 |
|  |  | No Data | 1 | 1 |
|  |  | % virus elimination | 70.6 | 46.7 |
|  |  | p (Fisher test ) | 0.2804 (ns) |  |
| 120h | ALL | Positives | 1 | 6 |
|  |  | Negatives | 23 | 17 |
|  |  | No Data | 6 | 10 |
|  |  | **% virus elimination** | **95.8** | **73.9** |
|  |  | p (Fisher test ) | **0.0479 (\*)** |  |
|  | **S** | Positives | 1 | 5 |
|  |  | Negatives | 11 | 7 |
|  |  | No Data |  | 4 |
|  |  | **% virus elimination** | **91.7** | **58.3** |
|  |  | p (Fisher test ) | **0.1550 (ns)** |  |
|  | **A** | Positives | 0 | 0 |
|  |  | Negatives | 12 | 10 |
|  |  | No Data | 6 | 6 |
|  |  | **% virus elimination** | **100** | **100** |
|  |  | p (Fisher test ) | **1.0000 (ns)** |  |

Medians times to reach SARS-CoV-2 elimination by RT-PCR were 3.0 and 5.0 days for the HeberFERON group and control group, respectively. A Kaplan-Meier plot of percent of SARS-CoV-2 positive patients along the first five days after treatment started showing statistical differences between groups through a Mantel Cox log-rank (p=0.0022) test (table 2 and figure 2). The Hazard Ratios of 3.2 and 95% CI of ratio of 1. 529 to 6.948 were calculated for the HeberFERON group as compared to the control group.

**Figure 2.**
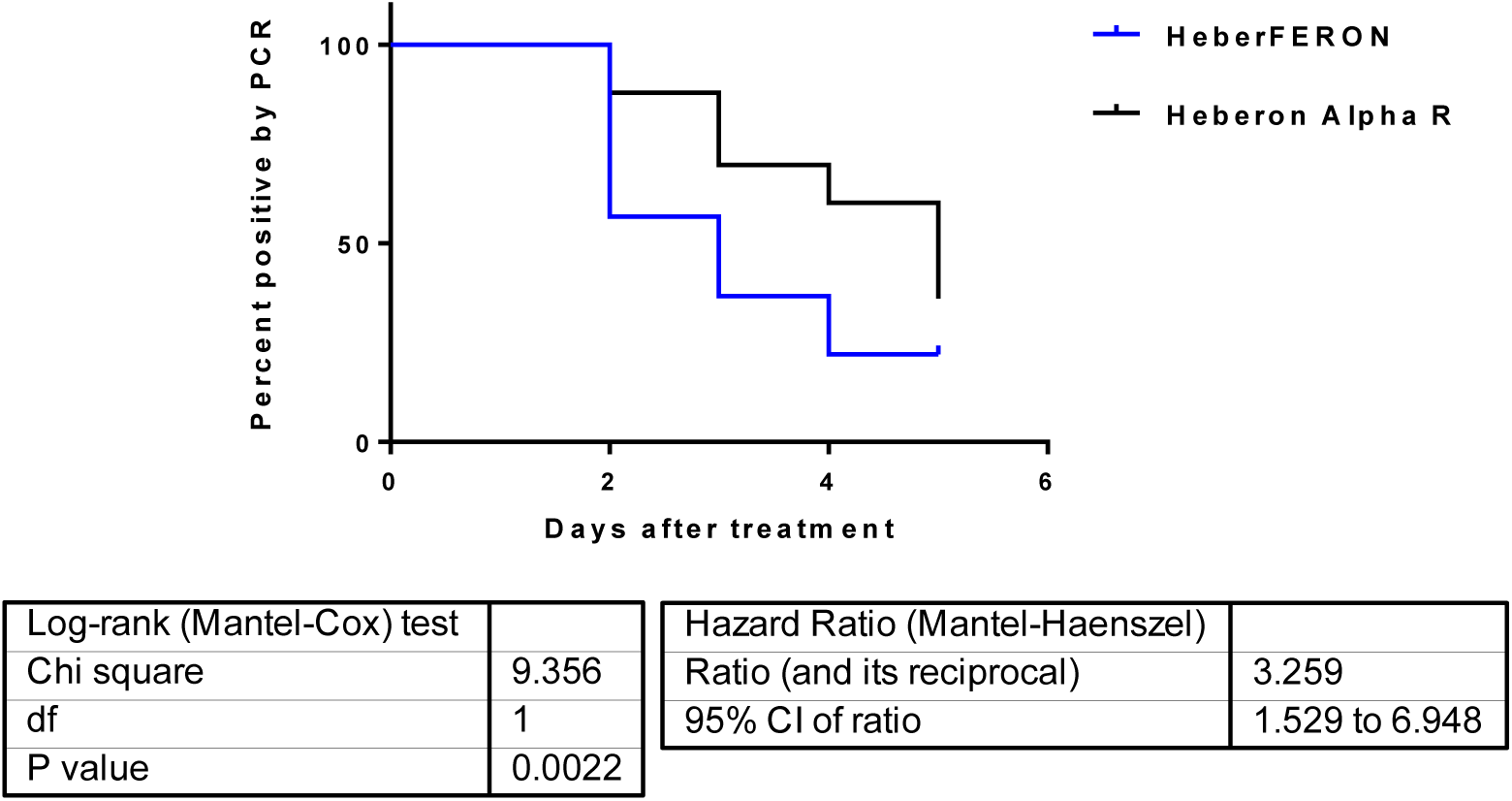
Kaplan-Meier representation for viral elimination. Viral elimination curves were constructed using GraphPrism (version 8) from the available viral detection data at four analysis points for the 63 patients split in two treatment groups. The P value (* p< 0.05) represents a statistical comparison of the two curves using a Log-rank Mantel-Cox test. Hazard Ratio (Mantel-Haenszel) and Median of viral elimination for the two treatments were also calculated.

When the viral RNA elimination was evaluated at 96 h after the treatment initiation, stratifying by the presence or absence of symptoms (see table 3), it was observed that 91.7% and 37.5% of symptomatic patients were negative for SARS-CoV-2 RNA in the HeberFERON and control groups, respectively, with statistical significance (p=0.006).

In asymptomatic patients, a lower rate of viral elimination was observed for both IFNs. However, the HeberFERON group showed a 70.6% of elimination in comparison to 46.7% for control group.

The worsening of respiratory symptoms was detected in two (6.6%) and one (3.3%) patients in HeberFERON and control groups, respectively but none of the patients advanced to severe COVID-19. The RT-PCR analysis after treatment with IFNs on day 14 (hospital discharge) was negative for SARS-CoV-2 in 100% and 91% of patients of HeberFERON and control cohorts, respectively.

Importantly, the kinetics for this recovery differed between treatment groups. The earlier increase in lymphocyte percentages was observed only for HeberFERON treated patients (p=0.0141) with a marked trend of an increment in lymphocyte concentrations. Also, a significant decrease in CRP (p= 0.0444) was observed for this group, parallel to a trend in the reduction in CPK (Figure 3).

**Figure 3.**
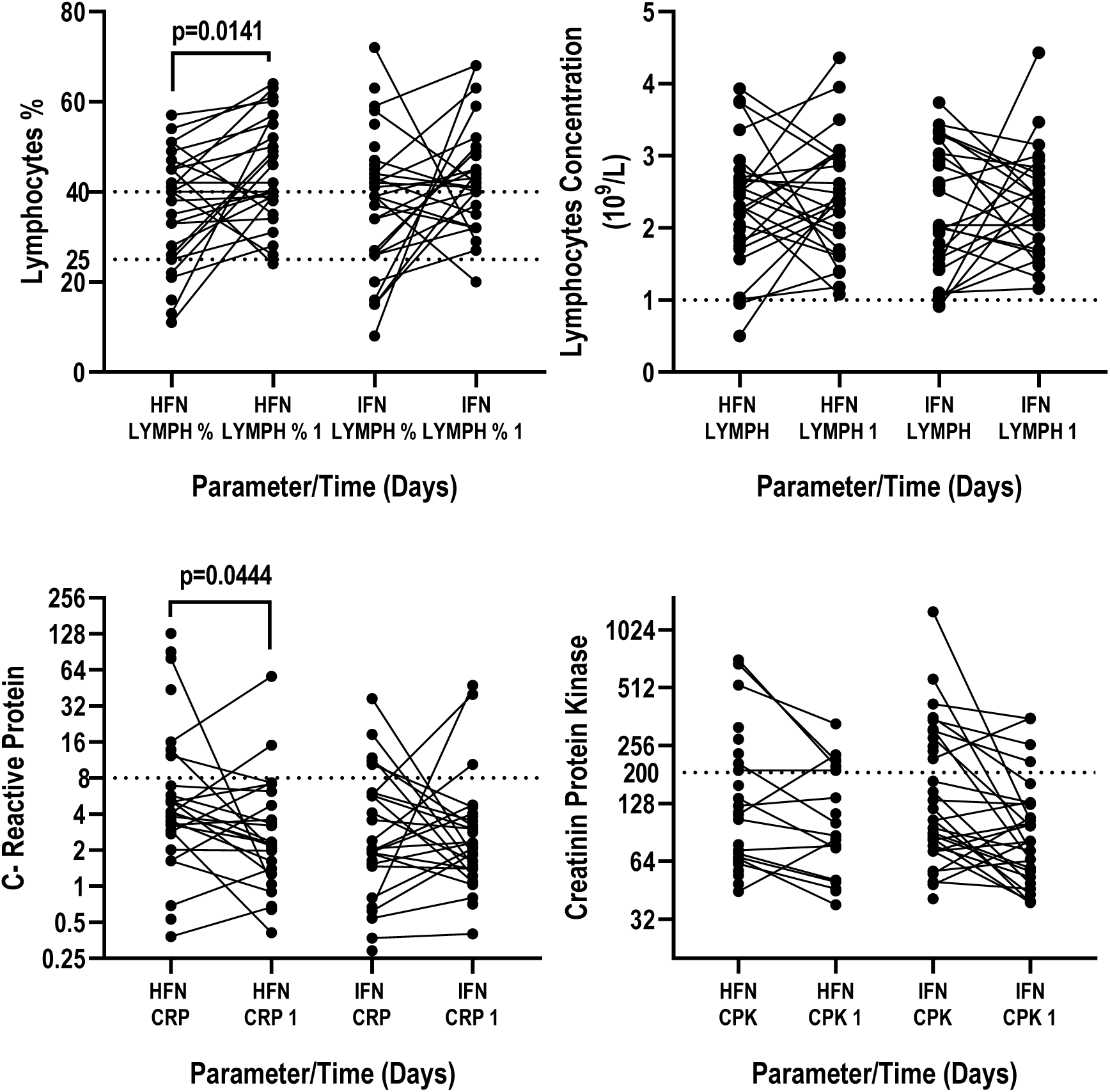
Comparative evaluation of clinical laboratory data a week after starting treatment in comparison to baseline. **(A)** Lymphocyte percentage among total leukocytes **(B)** Lymphocyte concentrations **(C)** Levels of C - reactive protein and **(D)** Creatine phosphokinase levels. Differences were analyzed across three time points, but only week one is shown. “P” values are indicated in cases of significant changes in the results of a mixed model adjustment for pair data sets. HFN: HeberFERON. LYMPH: Lymphocytes. CRP: C-reactive protein.

The correlation between clinical data evolution and SARS-CoV-2 virus clearance was analysed using a two-tailed non-parametric Spearman correlation with 95% confidence interval. Table 2 summarizes the parameters identified with significant direct or indirect relation with the reduction in the time needed to achieve a negative PCR result. A particular assessment of the same parameters is also included for the symptomatic patients included within each experimental group (Table 2). A statistically significant correlation between the viral elimination by HeberFERON on the first week of treatment was observed for lymphocyte concentration, C-reactive protein and the platelet to leukocyte ratio (PLR). Such a correlation was observed in the control group but only for CPK. Further stratification of the data on patients exhibiting symptoms or not exhibiting symptoms of the disease indicates that such correlations were maintained for lymphocyte concentration and PLR.

Adverse events were identified in 18 (31.5%) of 57 patients analyzed for such events, 8 (28.5%) in the control group and 10 (34.4%) in the HeberFERON group. The most common adverse events were headache (17.4%), nausea, hypertension, retroorbital pain and burning eyes (3.5%). Nighty-four percent of adverse events were mild and none were severe. There were no differences between the incidence of duration of any of the adverse events between the treatment groups. No serious adverse events were reported and no patients died during the study (Table 2).

## Discussion

Asymptomatic incubation period with or without detectable viral RNA, followed by non-severe symptomatic conditions and viral presence, ending in a severe symptomatic stage with high viral load, characterizes the SARS-CoV-2 infection^19^.

The global damage of COVID-19 could be partly explained by the median incubation time, from four to seven days to symptoms, a large window of time for transmission^20, 21^. Also, many infected patients remain completely asymptomatic and yet are fully capable of transmitting the virus^22,23,24^.

Insufficient activation of the IFN system is referred to as the principal cause of innate immune failure to control viral persistence. Importantly, an adaptive immune response is a fundamental factor needed for clearing and maintaining suppression of viral infections^25^.

The present study is the only randomized open-label controlled trial reported so far assessing the efficacy and safety of the combination of IFNs alpha-2b and gamma versus IFN-α2b in patients with COVID-19 ^26, 27^.

Very early (i.e. 48 h) after the first administration of HeberFERON, an important elimination of SARS-CoV-2 was obtained (45%); a result that is consistent with a potent and rapid antiviral effect. This fast response and the 96% viral elimination by day 5 of treatment with HeberFERON, has not been obtained for any other drug studied so far, even in a combinational design as described for the combined use of lopinavir 400 mg and ritonavir 100 mg every 12 h, ribavirin 400 mg every 12 h, and three doses of 8 MIU IFN beta-1b^14^. Even the 74% viral elimination seen with Herberon Alpha-2b is superior to the results reported by other authors^28^.

Time to reach the elimination of the SARS-CoV-2 as measured by RT-PCR was 3.0 and 5.0 days from the start of treatment with HeberFERON and Heberon alpha R, respectively, a difference that is statistically significant. These results are in concordance with in vitro data demonstrating the greater sensitivity of SARS-CoV-2 to IFNs^29, 30^ as compared to treatment with Heberon Alpha R alone.

Viral dissemination is an important determinant in the establishment of severe disease^31^. Therefore, the shortening of time to virus clearance as has been demonstrated for HeberFERON will impact very favorably in the disease outcome in COVID-19 patients.

The timing of initiation of antiviral therapy is another key factor in the treatment of viral infections. With respect to SARS-CoV, no effect of several antiviral drugs was observed when the treatments were started 6–14 days after symptom onset^32^. It was suggested that administration of antiviral medications at the beginning of the infection might improve outcome of patients with COVID-19^33^. Early treatment with IFNs was recommended in the treatment of MERS^34^ but late therapy (10–22 days)^32^ may contribute to poor outcomes^35^.

The Cuban Protocol for Management of COVID-19^2^ allowed us to include patients during a window of 7-8 days of symptoms onset. This is a good time point to start to reinforce the host innate and adaptive immune response with the use of IFNs, mainly with the combination of IFN-α2b and IFN-γ as this combination links the innate and adaptive immune response^36^. A recent open-label trial has demonstrated no benefit in hospitalized adult patients with severe COVID-19 treated with Kaletra. However, peer-protocol analyses suggested possible reductions in time to clinical improvement, particularly in those treated within 12 days of symptom onset^28^. The combination of Kaletra with other antiviral agents, as has been done in SARS^37^, MERS-CoV,^38^ and the trial reported here, might enhance antiviral effects and improve clinical outcomes. The confirmation of this therapeutic approach remains to be determined. However, it has been recently shown the combination of IFNs with Kaletra is associated with more favorable clinical outcomes than the use of Kaletra alone in COVID-19 patients^35^.

The presence of IFN-γ in the HeberFERON formulation, in additional to its strong immune regulatory functions, may restrict the angiotensin-converting enzyme 2 (ACE2) expression^39^, a receptor for SARS-CoV-2 cell entry^40^. It has been reported that this cytokine can directly inhibit viral entry for several viral infections (HCV and HIV) ^41^ by controlling the expression and/or distribution of their receptors. The effect of the combination of both IFNs on the ACE expression is under evaluation.

The results of virus elimination showed a more potent antiviral effect of HeberFRON over Heberon Alpha R alone, mainly in the symptomatic subjects. However, our study includes a low amount of symptomatic patients (46.0%) while in the control group there were more symptomatic patients (51.5%) than in the HeberFERON group (40.0%). Asymptomatic cohorts constitute the least manageable group of patients because they can spread the virus efficiently, as silent spreaders of SARS-CoV-2 which can cause difficulties in control of virus spread^22,42^. Symptomatic and asymptomatic patients can have similar viral loads^43^but the asymptomatic patients have a significantly longer duration of viral shedding as well as abnormal radiological findings in more than 90% of such patients^44^.

Although most countries do not pay adequate attention to asymptomatic people, this population of infected patients negatively impacts the global outcomes of the disease; therefore the treatment and follow-up of these patients are important to control the pandemic. The use of IFNs may be a determinant factor in the control of the disease in both symptomatic and asymptomatic patients, as reflected in this trial, where HeberFERON is most effective in the elimination of viral replication in both symptomatic and asymptomatic patients.

In MERS-CoV infected mice, delayed IFN treatment was associated with increased infiltration and activation of monocytes, macrophages, and neutrophils in the lungs; and enhanced pro-inflammatory cytokine expression^34^. Additionally, soon, after human infection, application of antiviral therapy with rapid viral clearance can delay pro-inflammatory cell development, activation and their infiltration, thus sparing patients of morbidity due to an overactive immune response^14^.

A delayed IFN response can also result in inflammation and tissue damage. The host may benefit from IFN presence early in the disease course, particularly if the IFN system is antagonized by viral proteins or is of a low response in older patients^24^.

An important difference between treatments concerns their effects on lymphocytes percentages within total leukocytes. Only in HeberFERON treated patients was a significant increase of lymphocytes percentages observed by week 1 and this finding, as well as lymphocyte concentrations, correlated significantly with the reduction in time to virus clearance (fig 3 and table 2).

It has been shown that lymphopenia predicts disease severity of COVID-19. In this context the restoration of lymphocyte population under the effect of a short and low dose of HeberFERON is very valuable. Further studies on the phenotype and functionality of peripheral blood mononuclear cell of patients are warranted since they will offer more clues with respect to the host response.

Although, IFNs have a potential for the induction of an inflammatory response, its early use contributes to regulate the initiation of the inflammatory response in COVID-19 patients^4^. However there have been only a few reports about the effect of IFN-α2b treatment in the reduction of the inflammatory mediators IL-6 and CRP in COVID-19 cases.^45^

The serum levels of CRP are not affected by factors such as age, sex, and physical condition, and correlate with the level of inflammation^46^. As such, this is an important index for the diagnosis and assessment of severe pulmonary infectious diseases^47,48^. In the early stage of COVID-19, CRP levels could reflect lung lesions and disease severity^49^as such levels correlate with the level of inflammation^46^. Herein we detected a significant reduction in CRP in patients after two administrations of HeberFERON (see figure 3). An important finding as CRP is an key index for the diagnosis and assessment of severe pulmonary infectious diseases^50,51^. In the early stage of COVID-19, CRP levels could reflect lung lesions and disease severity. The downregulation of CPR levels by IFNs in patients early in the disease could avoid acute inflammatory pathogenesis and disease severity^52^.

The administration of the HeberFERON in patients primed with the antivirals may result in a further boost of the host antiviral response, thus shortening the time for viral clearance and to lowering the probabilities of more severe disease development. This would certainly indicate that a lower lethality rate should occur as a result of this treatment.

Although with significantly more aged patients in the HeberFERON cohort (40% of symptomatic patients with a median age of 50 years-old), none of these patients became severely ill during the trial and all of them were eventually discharged. However, two patients from the HeberFERON group experienced worse respiratory symptoms. These were asymptomatic men at admission, both 33 years-old, with asthma as a comorbidity, that received 3 and 4 doses of HeberFERON. They were negative to SARS-CoV-2 RT-PCR at 48h and 96 h after the first HeberFERON administration. During the days of symptoms worsening, climate conditions were favorable for exacerbation of their asthma. Fortunately they recovered in 48 hours after anti-inflammatory therapy. In the control group a symptomatic man of 80 years-old, with hypertension as comorbidity, received only one dose of Heberon Alpha R and his respiratory symptoms worsened, requiring a transfer to the ICU. However, his RT-PCR for viral RNA was negative 20 days after symptoms had worsening and he was subsequently discharged.

Altogether these results indicate that with high probability, the rapid viral elimination detected in HeberFERON treated patients is translated into the reduction of systemic inflammation markers while inducing a significant increase in circulating lymphocyte concentrations. This increase may explain the symptomatic improvement observed in patients with high risk in the HeberFERON group. These results add to the anti-inflammatory effect described for IFNs in COVID-19 patients^45^, and are in agreement with the finding of of an IFN gene signature observed in mild-to-moderate COVID-19 patients^53^.

About 15% of the confirmed COVID-19 cases progress to the severe phase, with a higher risk for patients over 65 years-old^54^. Using this percentage, in the 63 patients included in our study, approximately 9 patients were expected to develop severe disease; however no patients became severely ill and no death occurred in these mild or moderate patients. In similar cohort of patients, 0.9% of mortality was described with the early use of IFNs^35^. In our trial several clinical parameters, known to be related to COVID-19 progression, were significantly improved by the treatments or showed a trend to a favorable outcome.

These results confirm the validity of early intervention with IFN treatment in patients with COVID-19. As demonstrated in the trial, the combination of type I and type II IFNs impacts strongly on the reduction of the risk for a patient with severe disease, likely through the efficient and timely implementation of a controlled inflammatory antiviral response against the SARS-CoV-2 infection.

HeberFERON formulation that combines in one vial IFN-α2b and IFN-γ results in an advantageous option for the treatment of COVID-19 patients. First, due to the demonstrated improved pharmacodynamics^16^ it is possible to administer less frequently and at lower doses than the other conventional IFNs, (IFN-α2b or IFN-β or IFN-λ). Furthermore, IFN-β has been used at doses 2 fold higher (of 12 MIU/mL^55^ or 8 MIU/mL^14^) when compared to HeberFERON doses. Second, the simultaneous administration of both types of IFNs will promote a faster and stronger innate and adaptive immune response and this synergy could be responsible for the quick clearance of SARS-CoV-2 as reported here.

It has been proposed that interferon is effective only in patients who lack comorbidities^56,57^; however we have obtained a high rate of viral elimination, resolution of symptoms, and full hospital discharge with HeberFERON treatment in a cohort of patients with 57% coexisting comorbidities. Moreover, it has been suggested that comorbidities, e.g. diabetes, affect the response to IFN^57^. However, two diabetic patients in our cohort eliminated the virus by day 3 and before day 14 of treatment.

Our study had several limitations. This trial was open label, without a placebo group with unbalanced demographics (age years) between treatment arms. In addition, sampling methods were most likely suboptimal using the throat sampling procedure, because of the inability to do sampling of lower respiratory tract secretions. Previous studies have shown that throat-swab specimens have lower viral loads^43^. Irrespective of these limitations the HeberFERON showed efficacy and was safe in shortening virus shedding, eliminating symptoms, and permitting discharge of patients with COVID-19.

Based in our analyses of the clinical data and their correlation with the patient outcome, we hypothesized that a potent antiviral response based in coordinated innate and adaptive immune responses was mediated by the combination of type I and type II IFNs. Thus an anti-inflammatory response in the early steps of the disease may be the main reason for the control of COVID-19 by HeberFERON treatment. The use of HeberFERON in COVID-19 patients could also be an effective therapeutic option to halt a second stage of the disease. This stage, characterized by a respiratory worsening approximately 9-12 days after onset of symptoms^58^, is apparently related to an imbalance of inflammatory mediators^53^.

Findings presented herein are the first to suggest therapeutic efficacy in COVID-19 disease of the combination of IFN-α2b and IFN-γ with individual and a public health impact, resulting in a shorter duration of virus shedding and preventing the worsening of the disease. Moreover, this is consistent with our understanding of the biology of IFNs and their synergistic response when used in combination^16^, that we have translated into a safe and effective antiviral therapy.

## Conclusion

HeberFERON was a safe treatment, superior to Heberon Alpha R in shortening the time to SARS-CoV-2 viral RNA elimination in a cohort of symptomatic or asymptomatic patients between 19 and 82 years-old, with more than 95% of patients negative for SARS-CoV-2 within 5 days of treatment.

The rapid viral elimination contributes to implement an anti-inflammatory response that can protect the patients from entering a more severe stage of the disease.

Early isolation combined with early administration of antiviral treatments, such as IFNs, is an efficient approach that could contribute to saving the life of many patients in the COVID-19 pandemic. Thus, the use of HeberFERON might be a distinctive choice in the preventive and therapeutic strategies necessary for current or future SARS outbreaks.

## Data Availability

The data are not available by now

